# Enhancing Semantic Interoperability in Precision Medicine: Converting OMOP CDM to Beacon v2 in the Spanish IMPaCT-Data Project

**DOI:** 10.1101/2024.12.25.24319606

**Authors:** Manuel Rueda, Juan Manuel Ramírez-Anguita, Victoria López-Sánchez, Sergi Aguiló-Castillo, Maria Eugenia Gas López, Alberto Labarga, Miguel-Ángel Mayer, Javier Ripoll Esteve, Ivo G. Gut

## Abstract

**Objective:** To introduce novel methods to convert OMOP CDM data into GA4GH Beacon v2 format, enhancing semantic interoperability within Spain’s IMPaCT-Data program for personalized medicine.

**Materials and Methods:** We utilized a file-based approach with the Convert-Pheno tool to transform OMOP CDM exports into Beacon v2 format. Additionally, we developed a direct connection from PostgreSQL OMOP CDM to the Beacon v2 API, enabling real-time data access without intermediary text files.

**Results:** We successfully converted OMOP CDM datasets from three research centers (CNAG, IIS La Fe, and HMar) to Beacon v2 format with nearly 100% data completeness. The direct connection approach improved data freshness and adaptability for dynamic environments.

**Discussion and Conclusion:** This study introduces two methodologies for integrating OMOP CDM data with Beacon v2, offering performance optimization or real-time access. These methodologies can be adopted by other centers to enhance interoperability and collaboration in health data sharing.

## 1. Introduction

Achieving semantic interoperability in health data is crucial for advancing precision medicine, particularly in handling the ever-growing scale and complexity of genomic and clinical datasets [1, 2]. In this regard, the OMOP Common Data Model (CDM) is a widely adopted framework for harmonizing clinical data, powered by the OHDSI standardized vocabulary [3]. However, its design lacks native support for genomic data, a critical component in personalized medicine.

To address this limitation, the OHDSI community has established a dedicated working group focused on expanding OMOP CDM to support genomic data integration (see https://github.com/OHDSI/Genomic-CDM). These efforts align with parallel initiatives such as the Global Alliance for Genomics and Health (GA4GH), which introduced Beacon v2 [4] [4] and Phenopacket v2 [5] standards for federated genomic and phenotypic data sharing [6, 7].

Rather than replacing OMOP CDM, the goal of GA4GH standards is to provide structured, interoperable data schemas for seamless integration across systems.

The Spanish National Infrastructure for Precision Medicine (IMPaCT) program aims to embed precision medicine into the Spanish National Health System, ensuring equitable access to cutting-edge genomic healthcare (https://impact.isciii.es/). Within this, the IMPaCT-Data initiative consolidates diverse datasets, leveraging Beacon v2 to enable robust genomic and phenotypic data discovery (https://impact-data.bsc.es/en/about/impact).

This article outlines the conversion of OMOP CDM data to Beacon v2 within the IMPaCT program. We introduce two methodologies: (i) a file-based approach for rapid data retrieval and (ii) an on-the-fly approach enabling real-time access. By addressing challenges encountered in real-world data transformation, this study offers actionable insights and practical solutions for advancing federated health data interoperability.

## 2. Materials and methods

The goal of this study is to enable researchers to query OMOP CDM patient data via the Beacon v2 API (see Figure 1), particularly for identifying individuals with specific characteristics or phenotypes. To achieve this, we implemented and evaluated two approaches: i) a file-based conversion, leveraging preformatted JSON data stored in a non-relational database like MongoDB, and, ii) an on-the-fly conversion, dynamically converting data from SQL-based relational databases (e.g., PostgreSQL) in real-time. These methods address different needs, such as efficient pre-processed data retrieval and real-time data access.

**Figure 1:**
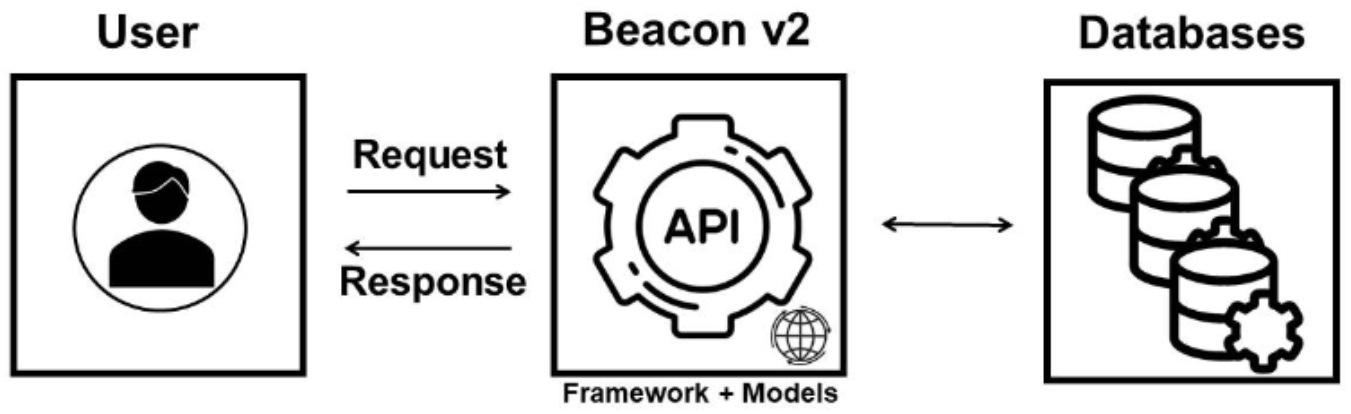
Schematic representation of how the Beacon v2 API functions.

### 2.1 File-based conversion (Convert-Pheno)

OMOP CDM databases often store millions of rows of patient data, making complex, multi-table queries computationally intensive. To address this, we pre-formatted OMOP data into the Beacon v2 exchange format, the Beacon Friendly Format (BFF), which is specifically designed to facilitate efficient data sharing and interoperability [8].

The process begins by exporting PostgreSQL data as text files (.SQL or .CSV), which are then transformed into JSON using Convert-Pheno [9], a tool developed under the 3TR project (https://3tr-imi.eu/) to convert phenotypic and clinical data into the Beacon v2 schema (see Supporting Text 1). The conversion process is typically performed once, with updates applied as new data becomes available. A tutorial for this file-based approach is available on a Google Colab document (https://github.com/CNAG-Biomedical-Informatics/omop-cdm-2-beacon-v2).

Once generated, the BFF JSON file can be loaded as a schema-validated collection into a non-relational database, such as MongoDB. These collections are readily accessible through the ELIXIR Beacon v2 Reference Implementation (B2RI) [8] or other Beacon v2 APIs based on non-relational databases (see Supporting Text 1 and Supporting Figure 1).

### 2.2 ‘On-the-fly’ conversion (Beacon v2 RI OMOP extension)

An alternative method involves directly linking the Beacon v2 API with the OMOP CDM database (see Figures 2 and 3), a technique implemented by members of the Barcelona Supercomputing Center (BSC) as an extension of the B2RI (https://gitlab.bsc.es/impact-data/impd-beacon_omopcdm) [10].

**Figure 2:**
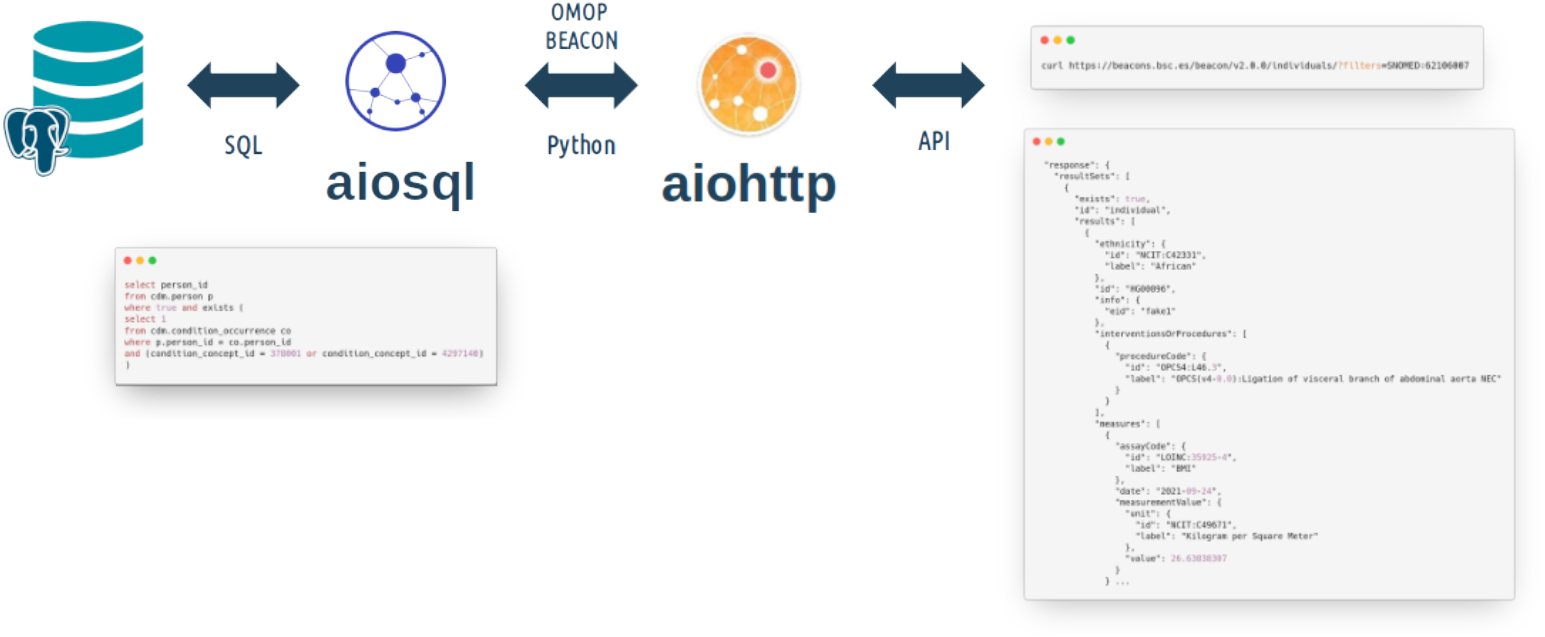
Current implementation of the Beacon v2 RI OMOP extension.

**Figure 3:**
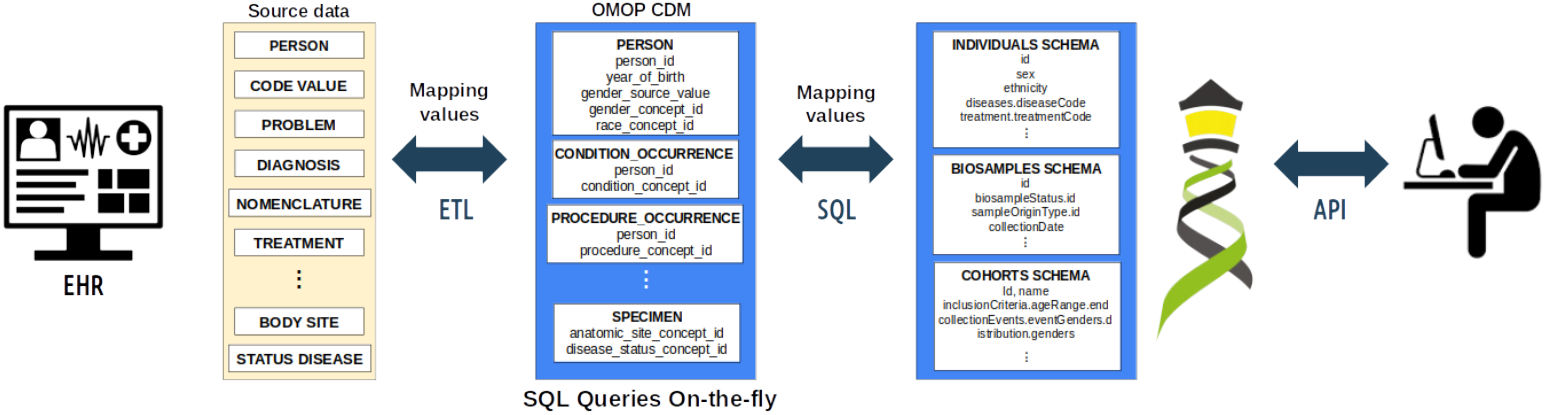
Schematic representation of the Beacon v2 RI OMOP extension.

This ‘on-the-fly’ approach involves connecting the Beacon v2 API directly to the OMOP CDM database using SQL queries (see Figure 3). The system translates the Beacon API requests into SQL queries that retrieve the necessary data directly from the database and transforms the data to JSON. This approach eliminates the need for the intermediate data exchange files, periodic transformations and the maintenance of the MongoDB replica of the data, providing an up-to-date view of the data with high degree of granularity.

## 3. Results

This section details the conversion of three OMOP CDM instances to the Beacon v2 format, conducted as part of Spain’s IMPaCT-Data project (GdT05-03). We used diverse clinical research databases, including a small dataset from the Centro Nacional de Análisis Genómico (CNAG) in Barcelona, a medium-scale dataset from the Health Research Institute Hospital La Fe (IIS La Fe) in Valencia, and a large dataset from Hospital del Mar (HMar) in Barcelona (see full details in Supporting Table 3).

### 3.1 File-based conversion at CNAG (with synthetic data)

At CNAG, we evaluated the file-based approach using the synthetic EUNOMIA dataset (https://ohdsi.github.io/Eunomia/), which includes records for 2,694 patients across multiple tables. This dataset was selected to validate the conversion process under controlled conditions. Data were loaded into PostgreSQL using R and exported as an .SQL file for transformation (see Table 1 and Supporting Table ST2 for details). Scripts are available upon request.

**Table 1:** Completeness on the conversion from OMOP CDM tables to Beacon v2 objects for the EUNOMIA synthetic dataset.

| OMOP tables | BFF term | Rows in CSV | Objects in BFF | Completeness (%) |
| --- | --- | --- | --- | --- |
| Concept | - | 444 | - | - |
| Condition_occurrence | diseases | 65,332 | 65,332 | 100 |
| Drug_exposure | treatment | 67,707 | 67,707 | 100 |
| Measurement | measures | 44,053 | 44,053 | 100 |
| Observation | phenotypicFeatures | 1,477 | 1,477 | 100 |
| Person | id | 2,694 | 2,694 | 100 |
| Procedure_occurrence | interventionsOrProcedures | 37,409 | 37,409 | 100 |
| Visit_occurrence | _visit | 1,037 | - | - |

Table 1 summarizes the completeness of converting EUNOMIA table records to the Beacon v2 “individuals” entity. VISIT_OCCURRENCE values are excluded because visits may span multiple terms per individual (stored under the “_visit” property). Additionally, handling longitudinal data is beyond the scope of the Beacon v2 specification.

### 3.2 File-based conversion at IIS La Fe

The conversion process from an OMOP CDM instance to the *individuals* entity of the Beacon v2 Models was executed using real data from the Big Data Platform of the IIS La Fe. Specifically, a retrospective study was conducted using pseudo-anonymised clinical information collected from patients (both survivors and non-survivors) infected with COVID-19 between December 31, 2021 and February 28, 2022 with a confirmed diagnosis of COVID-19 via RT-qPCR.

Clinical information was extracted from various sources, including pharmaceutical prescriptions, symptomatology, clinical observations, hematology, biochemical and blood gas parameters, and hospital and primary care resources, all mapped to OMOP CDM. The resulting COVID-19 dataset encompassed records for 18,554 patients across several tables: CONCEPT, CONDITION_OCCURRENCE, DRUG_EXPOSURE, MEASUREMENT, OBSERVATION, PERSON, PROCEDURE_OCCURRENCE, and VISIT_OCCURRENCE. Each aforementioned table was exported from SQL databases to 8 CSV files. The Convert-Pheno command line used was: ‘convert-pheno -iomop *.csv -obff individuals.json’ (see details in Supporting Table ST2). The table 2 illustrates the result of the transformation of COVID-19 records into the *individuals* entity.

**Table 2:**
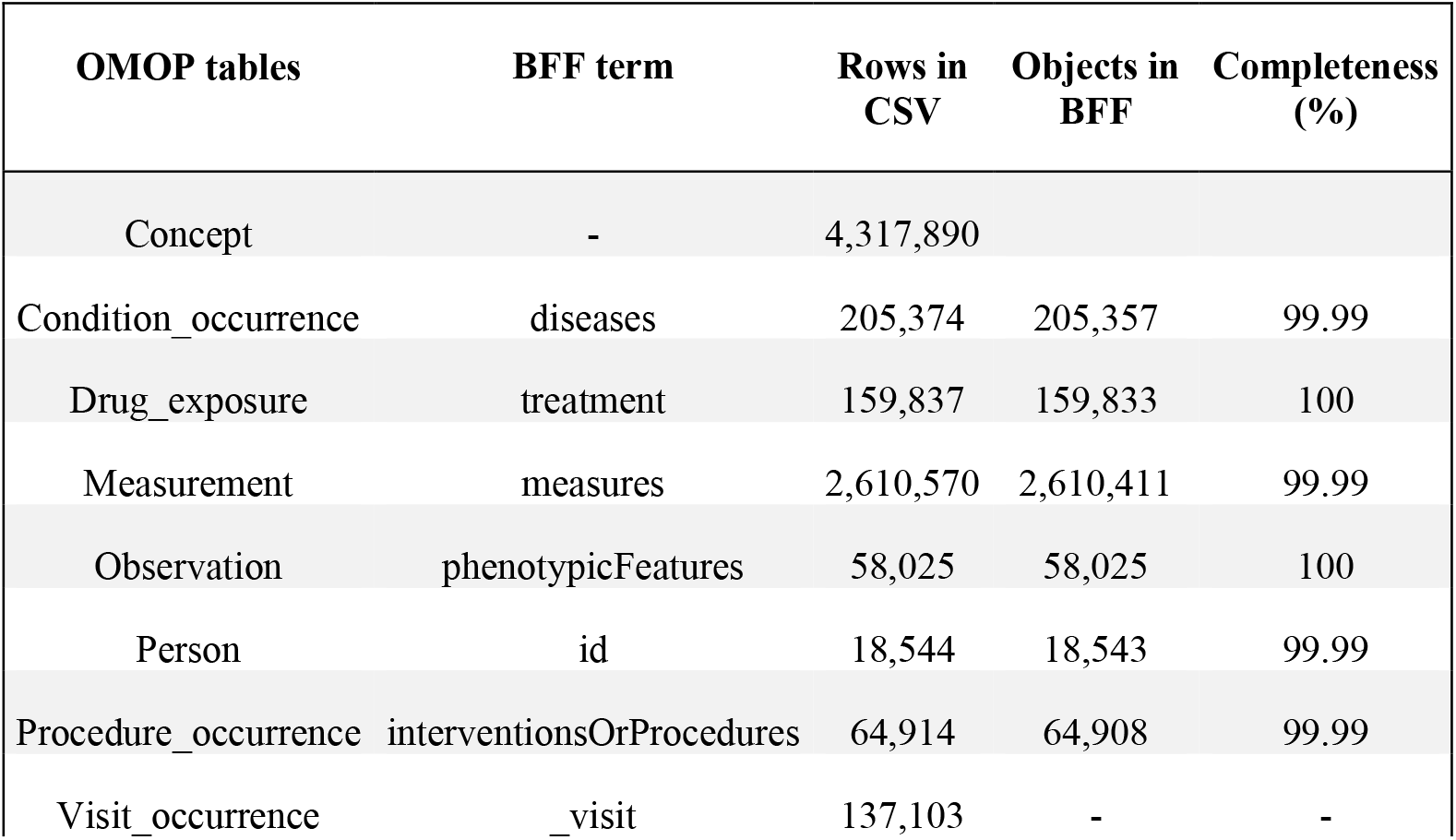
Completeness on the conversion from OMOP CDM tables to Beacon v2 objects for the COVID-19 dataset.

### 3.3 File-based conversion at HMar

The data conversion process begins with the IMASIS information system, the EHR system of Parc Salut Mar Barcelona, a comprehensive healthcare services organization. Currently, this information system includes and shares the clinical data of two general hospitals, one mental health care centre, one social-healthcare centre and five emergency room settings, which are offering different services in the Barcelona city area (Spain). IMASIS includes clinical information from around 1 million patients who have used the services of this healthcare system since 1990 and from different settings such as admissions, outpatients, emergency rooms, and major ambulatory surgery. The diagnoses are coded using The International Classification of Diseases ICD-9-CM [11] and ICD-10-CM [12]. IMASIS-2 is the anonymized relational database of IMASIS being the data source used for mapping to OMOP including additional sources of information such as the Tumours Registry.

The IMASIS-2 OMOP dump file was used as input for Convert-Pheno in ‘stream’ mode with the command: ‘convert-pheno -iomop dump.sql -obff individuals.json -stream -max-lines-sql 999999999’. As shown in Table 3, after the conversion, the completeness of the mappings approaches 100% across all domains.

**Table 3:** Completeness on the conversion from OMOP CDM tables to Beacon v2 objects for the IMASIS-2 dataset.

| OMOP tables | BFF term | Rows in CSV | Objects in BFF | Completeness (%) |
| --- | --- | --- | --- | --- |
| Concept | - | 6,871,345 | - | - |
| Condition_occurrence | diseases | 7,837,107 | 7,837,107 | 100 |
| Drug_exposure | treatment | 73,400,506 | 73,400,506 | 100 |
| Measurement | measures | 248,897,994 | 248,885,056 | 99.99 |
| Observation | phenotypicFeatures | 1,672,053 | 1,672,053 | 100 |
| Person | id | 1,033,805 | 1,033,623 | 99.98 |
| Procedure_occurrence | interventionsOrProcedures | 1,815,641 | 1,815,641 | 100 |
| Visit_occurrence** | _visit | 16,270,148 | - | - |

\*\*Table VISIT\_OCCURRENCE was excluded from the calculation.

### 3.4 ‘On-the-fly’ conversion

The novel ‘on-the-fly’ conversion method is a promising alternative for real-time data access and conversion. Early tests indicated great efficiency and robustness, particularly in scenarios requiring immediate data access. Within the IMPaCT project, this approach is currently being developed in the context of the UNICAS IMPaCT pilot project (https://www.sjdhospitalbarcelona.org/es/hospital/proyectos-estrategicos/red-unicas-atencion-enfermedades-minoritarias), to create a Federated Pediatric Network for Personalized Medicine in Rare Pediatric Diseases led by Hospital Sant Joan de Deu (Barcelona).

## 4. Discussion

This study is the first to explore converting OMOP CDM data to Beacon v2 in a real-world setting, complementing prior work on integrating OMOP with GA4GH standards like Phenopackets v2 [13, 14] [15]. The following sections outline the advantages of the two conversion approaches—file-based and on-the-fly—in different operational contexts (see also Table 4).

**Table 4:** Comparison of file-based and ‘on-the-fly’ OMOP CDM to Beacon v2 conversion approaches.

|  | File-Based Conversion | Direct SQL Query |
| --- | --- | --- |
| **Data Freshness** | Data are static and requires manual updates | Real-time data access, ensuring the latest information is always available |
| **Integration with Genomic Data** | Unified Backend: Uses MongoDB to centralize clinical and genomic data (from VCFs). | Database Duplicity: Separate databases for genomic data (MongoDB) and clinical data (SQL-based) |
| **Performance** | Fast response with pre-formatted data | Fast response with properly indexed OMOP database |
| **Implementation Complexity** | Requires software installation and a secure environment to process data | Simplified setup by avoiding intermediate steps, directly querying the database |
| **Resource Utilization** | Requires storage proportional to data size. Data must be ingested and indexed into a non-relational database | Efficient use of storage as data remains in the database |
| **Scalability** | Scalable with pre-formatted (JSON Schema validated) data | Dynamically scalable with efficient SQL queries and robust and mature SQL technologies |
| **Flexibility** | Limited to Beacon v2 data schema | Highly flexible with dynamic queries |
| **Maintenance** | May require periodic updates | Lower maintenance with automatic data freshness |
| **Data Security** | Controlled data dumps disconnect Beacon v2 API requests from OMOP instances, adding a layer of security. Text files still require secure handling. | Enhanced security by keeping data within the secured database environment |
| **Data Reusability and FAIRification\*\*** | Converted data can be directly deposited into repositories such as the EGA [17] or stored in data warehouses from EU-funded projects. Data are immediately available for analytics (e.g. [19]) | Directly relies on open source OMOP tools like Atlas or HADES for flexible, FAIR-aligned data reuse and analysis |

1 \*\* Findable, Accessible, Interoperable, Reusable

### When to Use File-Based Conversion

The file-based approach is well-suited for centers leveraging non-relational databases, such as MongoDB, to manage genomic data derived from VCFs [16] alongside clinical data. It is particularly effective for projects consolidating data from multiple sources (e.g., REDCap, CDISC, OMOP CDM, CSV) into centralized storage, enabling standardized data sharing through the Beacon v2 API. A key advantage of this approach is the fast API response times achieved by preformatted data, making it ideal for workflows where data transformation occurs once data collection is complete—similar to submissions to repositories like EGA [17] and dbGAP [18], which mandate the simultaneous processing of both genomic and clinical data. However, this method requires periodic exports to reflect new data, which may be a limitation in dynamic environments. Despite this, the method is computationally efficient, compatible with standard server environments, and secure when conducted in controlled settings that safeguard data sensitivity and anonymity

### When to use ‘On-the-fly’ Conversion

The on-the-fly conversion approach is beneficial when access to the most up-to-date information in the OMOP CDM is essential (e.g., updates to OMOP CDM are frequent). It also eliminates the need for periodic data transformation and avoids maintaining duplicate clinical data, thereby reducing the demand for additional computational resources. However, it requires a well-indexed and optimized database to support complex SQL queries efficiently and it is not ideal for scenarios requiring a unified backend with genomic data stored in a non-relational database (e.g., ingested BFF from processed VCFs). In addition to the ongoing example within the IMPaCT project (See Methods) the Beacon v2 RI OMOP extension is already being used for the COVID-NL dataset in the By-Covid project (https://by-covid.org/). It is also planned to deploy the implementation in the DATOS-CAT project, which will provide access to the Spanish GCAT cohort data (http://www.gcatbiobank.org/).

## 5. Conclusion

This study introduces two complementary approaches for transforming OMOP CDM data into the Beacon v2 schema: a file-based method for efficient preprocessed data retrieval and an on-the-fly conversion for real-time access. By offering practical guidance, we aim to support similar initiatives and encourage the broader adoption of Beacon v2.

These methodologies improve the interoperability and accessibility of clinical and genomic data, fostering collaboration and advancing precision medicine to enhance patient care.

## Data Availability

All data produced in the present study are available upon reasonable request to the authors.

## Acknowledgements

We would like to express our gratitude to the current and past members of the EGA, particularly Amy Curwin for logistical support and Babita Singh and Jordi Rambla for their initial feedback. Special thanks to all members of the GdT3-05 group within the IMPaCT-Data project. We extend our appreciation to Salvador Capella-Gutiérrez for his valuable assistance in reviewing this manuscript.

## Ethics approval and consent to participate

Health Research Institute Hospital La Fe: All data extraction methods adhered to relevant guidelines and regulations, with the study receiving approval on May 29, 2024 from the Ethics Committee for Biomedical Research with Medicines of the University and Polytechnic La Fe Hospital of Valencia, under registration number 2024-0484-1. IMASIS-2 was used with permission of the Ethics Committee of the Hospital del Mar, under registration number 2023/11262.

## Declaration of generative AI and AI-assisted technologies in the writing process

During the preparation of this work the author(s) used ChatGPT 4 by OpenAI to improve readability and language of some of the paragraphs. After using this tool/service, the author(s) reviewed and edited the content as needed and take(s) full responsibility for the content of the publication.

