## Supplementary Data for "Enhancing Semantic Interoperability in Precision Medicine: Converting OMOP CDM to Beacon v2 in the Spanish IMPaCT-Data Project"

### Sup. Data Contents:

BFF integration with the Beacon v2 API ........ STxt1 and SF1
Mapping between OMOP CDM and Beacon v2 schemas ........ STxt2 and ST1
Comparative Performance Metrics for Data Conversion Across Different Centers........ ST2
References

### Supporting Text 1 and Figure 1: BFF integration with the Beacon v2 API

This work focused on integrating phenotypic and clinical data from the OMOP CDM schema into Beacon v2, a standard for genomic and phenotypic data sharing. Specifically, the integration targeted the *individuals* entity of the Beacon v2 Models (https://docs.genomebeacons.org/), which includes terms such as *diseases*, *ethnicity*, *exposures*, *geographicOrigin*, *id*, *info*, *interventionsOrProcedures*, *karyotypiSex*, *measures*, *pedigrees*, *phenotypicFeatures*, *sex*, and *treatments*. These elements capture both patient and healthy control data, supporting a wide range of use cases in precision medicine.

The workflow involves creating a JSON output file (e.g., 'individuals.json') using the Convert-Pheno tool [1]. This was achieved through the command-line interface (CLI) with the default command: 'convert-pheno -iomop file.sql -obff individuals.json -max-lines-sql 999999'. By default, Convert-Pheno operates in 'no-stream' mode, loading all patient data from SQL tables into RAM, transposing and merging rows into single patient objects (details in Supporting Text 2 and Supporting Table ST1). For memory-limited environments, the 'stream' mode processes OMOP tables row by row, using the term *id* as primary key to avoid loading all data into memory. During conversion, "concept_id" values in the CONCEPT table remain unchanged, except for the *sex* term, which is mapped to NCIt terminology. Necessary "concept_id" entries are included to maintain schema consistency.

This 'invididuals.jon' file serves as the foundation for data ingestion into a MongoDB instance, which acts as the backend for the Beacon v2 Reference Implementation API (*beacon2-ri-api*) [2]. Figure SF1 illustrates the end-to-end process, from OMOP CDM data extraction to Beacon v2-compatible JSON generation and MongoDB integration.

In this setup, the MongoDB instance is deployed in a dedicated Docker container for scalability and isolation. The *beacon2-ri-api* was initially chosen for its compatibility with BFF-based MongoDB workflows, with deployment documentation available at https://github.com/EGA-archive/beacon2-ri-api. Alternative APIs, such as BSC's Java-based implementation (https://github.com/ga4gh-beacon/java-beacon-v2) [3] or the newest *beacon2-pi-api* (https://github.com/EGA-archive/beacon2-pi-api), can also be used. This approach ensures efficient, scalable, and standardized data sharing while maintaining compatibility with the Beacon v2 ecosystem and adhering to FAIR data principles (Findable, Accessible, Interoperable, Reusable).

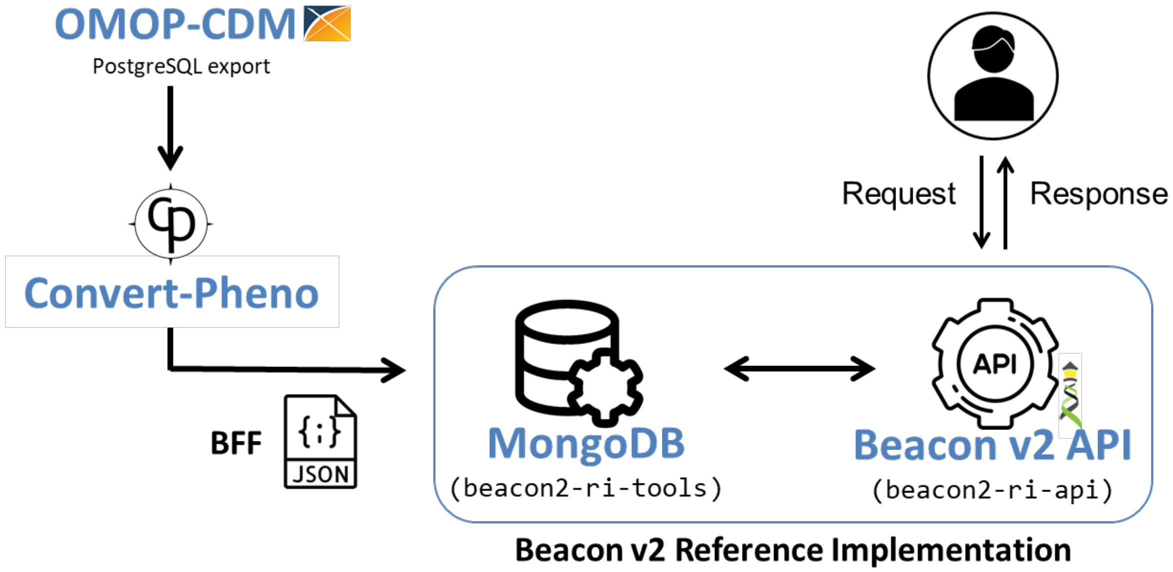


**Figure SF1:** Overall flow of the file-based OMOP CDM to Beacon v2 conversion. Adapted from the Convert-Pheno publication [1] with additional information.

### Supporting Text 2 and Table 1: Mapping between OMOP CDM and Beacon v2 schemas

The OMOP Common Data Model (CDM) v5.4 employs a relational SQL schema (https://ohdsi.github.io/CommonDataModel/cdm54.html), whereas Beacon v2 adopts a hierarchical JSON schema (https://docs.genomebeacons.org/schemas-md/individuals_defaultSchema). Bridging these formats requires a systematic transition from a table-based structure to a hierarchical representation, ensuring compatibility without data loss.

To address this, schema mappings were developed during the creation of the Convert-Pheno tool [1]. These mappings align the relevant OMOP CDM tables—such as CONCEPT, CONDITION_OCCURRENCE, DRUG_EXPOSURE, MEASUREMENT, PERSON, PROCEDURE_OCCURRENCE, OBSERVATION, and VISIT_OCCURRENCE—with corresponding elements in the Beacon v2 *individuals* default schema. The latest mappings and step-by-step instructions are available in the online documentation at [https://cnag-biomedical-informatics.github.io/convert-pheno/omop2bff/](https://cnag-biomedical-informatics.github.io/convert-pheno/omop2bff/).

Key Considerations in Mapping:

1. Lossless Data Transition: To ensure no critical data is lost, all unmapped OMOP variables are stored under the "info" term in the Beacon v2 schema. This practice safeguards data completeness while maintaining compatibility.
2. Custom Selection of Exposures: For the Beacon v2 *exposures* term, a manual selection of commonly used 'concept_id' values was made from the OMOP

CONCEPT table. Details of the selected concepts can be found at https://github.com/CNAG-Biomedical-Informatics/convert-pheno/blob/main/share/db/concepts_candidates_2_exposure.csv.

Process Overview:

The mapping process bridges the structural gap between the two models, translating complex OMOP relational data into a semantically rich and hierarchically organized Beacon v2 format. This ensures that clinical and phenotypic information is preserved and accessible for advanced genomic and precision medicine workflows.

See examples of the mapping at Supporting Table 1. For detailed step-by-step mapping instructions, refer to https://cnag-biomedical-informatics.github.io/convert-pheno/mapping-steps.

**Supporting Table 1**: Example of Beacon v2 *individuals* entity terms mapped from clinical events in OMOP CDM.

| Beacon Term (Individuals) | BFF (output) | OMOP "Fact table" | OMOP mapping based on | |
|---|---|---|---|---|
| **ethnicity** | **"ethnicity": {**<br>"id": "NCIT:C41261",<br>"label": "White"<br>} | person | concept_id | 8527 |
| | | | domain_id | race |
| | | | concept_name | White |
| **id** | **"id"**: "1" | person | person_id | 1 |
| **sex** | **"sex"**: {<br>"id": "NCIT:C20197",<br>"label": "Male"<br>} | person | gender_concept_id | 8507 |
| | | | domain_id | gender |

| | | | | |
|---|---|---|---|---|
| **info** | **"info"**: {<br>"PERSON": {<br>"OMOP_columns": {<br>(…) }<br>}, | person | | |
| **diseases** | **"diseases"**: [<br>{<br>"_info": {<br>"CONDITION_OCCURRENCE": {<br>"OMOP_columns": {<br>(…) }<br>},<br>"ageOfOnset": {<br>"age": {<br>"iso8601duration": "32Y"<br>}<br>},<br>"diseaseCode": {<br>"id": "SNOMED:444470001",<br>"label": "Injury of anterior cruciate ligament"<br>} | condition_occurrence | condition_concept_id | 40479768 |
| | | | domain_id | condition |
| | | | concept_name | Injury of anterior cruciate ligament |
| | | | condition_start_date | ‘’ |
| | | | birth_date time (person) | 1949-01-27 00:00:00.000 |
| | | | vocabulary_id | SNOMED |
| | | | concept_code | 444470001 |
| **interventionsOr Procedures** | **"interventionsOrProcedures"**: [<br>{<br>"_info": {<br>"PROCEDURE_OCCURRENCE": {<br>"OMOP_columns": {<br>(...) }<br>},<br>"ageAtProcedure": {<br>"age": {<br>"iso8601duration":"32Y"<br>}<br>},<br>"dateOfProcedure": "1981-08-17",<br>"procedureCode": {<br>"id": "SNOMED:699253003",<br>"label": "Surgical manipulation of joint of knee"<br>} | procedure_occurrence | procedure_concept_id | 44783196 |
| | | | domain_id | procedure |
| | | | concept_name | Surgical manipulation of joint of knee" |
| | | | procedure_start_date | ‘’ |
| | | | birth_date time (person) | 1949-01-27 00:00:00.000 |
| | | | vocabulary_id | SNOMED |
| | | | concept_code | 699253003 |

<table>
<tr><td rowspan="7">treatments</td><td rowspan="7">"treatments": [<br>{<br>"_info": {<br>"DRUG_EXPOSURE": {<br>"OMOP_columns": {<br>(…) }<br>},<br>"ageAtOnset": {<br>"age": {<br>"iso8601duration":<br>"36Y"<br>}<br>},<br>"doseIntervals": [],<br>"routeOfAdministration":<br>{<br>"id": "NCIT:N0000",<br>"label": ""<br>},<br>"treatmentCode": {<br>"id": "RxNorm:140587",<br>"label": "celecoxib"<br>}</td><td rowspan="7">drug_exposure</td><td>drug_concept_id</td><td>1118084</td></tr>
<tr><td>domain_id</td><td>drug</td></tr>
<tr><td>concept_name</td><td>celecoxib</td></tr>
<tr><td>drug_exposure_start_da<br>te</td><td>‘’</td></tr>
<tr><td>birth_date time (person)</td><td>1949-01-27<br>00:00:00.000</td></tr>
<tr><td>vocabulary_id</td><td>RxNorm</td></tr>
<tr><td>concept_code</td><td>140587</td></tr>
</table>

### Supporting Table 2: Comparative Performance Metrics for Data Conversion Across Different Centers

| | CNAG | IIS La Fe | HMar |
|---|---|---|---|
| **Data Type / Name** | Synthetic / EUNOMIA | Real / COVID-19 | Real / IMASIS 2 |
| **# Patients** | 2,694 | 18,554 | 1,033,805 |
| **# Records** | ~220 K | ~8 M | ~350 M |
| **Timing / Hardware**** | ~5 sec / Intel® Xeon® W-1350P CPU @ 4.00GHz with 32GB RAM | < 4 min / Intel® Core™ i9-13900K with 125 GB RAM | 7.1 hours / Intel® Xeon® CPU W-2265 @ 3.50GHz with 188 GB RAM |
| **Input Type / Size** | PostgreSQL export / 33 MB (7MB gz) | CSVs / 944 MB (127 MB .tar.gz) | PostgreSQL export / 56 GB (8.1 GB gz). The dump creation took ~6 minutes. |
| **Output (JSON array) Size** | 2,694 objects / 354 MB (19 MB gz) | 18,544 objects/ 6 GB (225 MB gz) | 333.6 million objects / 1.2TB (19 GB gz) |
| **Mode / RAM Usage** | No-stream / ~500 MB | No-stream / ~15 GB | Stream / ~8 GB |

** 1 core. Further information on system requirements can be found at the online documentation (https://cnag-biomedical-informatics.github.io/convert-pheno/omop-cdm/#omop-as-input).
